# Determinants of Bluetooth-enabled Self Measured Blood Pressure monitoring in Federally Qualified Health Centers

**DOI:** 10.1101/2023.08.17.23294249

**Authors:** Abby Hellem, Candace Whitfield, Maria Mansour, Yvonne Curran, Mackenzie Dinh, Kimberly Warden, Lesli Skolarus

**Affiliations:** Department of Neurology, Michigan Medicine, University of Michigan, Ann Arbor, MI; Emergency Medicine, Michigan Medicine, University of Michigan, Ann Arbor, MI; Davee Department of Neurology, Northwestern University, Chicago, IL; Hamilton Community Health Network, Flint, MI

**Keywords:** Hypertension, mhealth, health equity, community, qualitative

## Abstract

**Background:** In 2021, the Health Resources and Services Administration (HRSA) launched the National Hypertension Control Initiative (HTN Initiative) with the goal to enhance HTN control through Bluetooth-enabled self-measured blood pressure (BT SMBP) monitoring and use this data to inform clinical decisions in Federally Qualified Health Centers (FQHCs) with large proportion of their population with uncontrolled BP. We sought to understand the experience of Michigan-based FQHCs in implementing the HTN initiative.

**Methods:** Staff from three Michigan-based FQHCs were invited to participate in semi-structured interviews from September to November 2022. Interviews were conducted in-person and were based on the Tailored Implementation of Chronic Diseases framework. Content analysis was performed by three coders.

**Results:** Ten staff participated in interviews (FQHC 1: n=6, FQHC 2: n=1, FQHC 3: n=3). The FQHCs differed in their stage of implementation and their approach. FQHC 1 created a large-scale, community health worker driven program, FQHC 2 created a small-scale, short term, BP device loan program, and FQHC 3 created a primarily outsourced, large-scale program through a contracted partner. Positive staff attitudes and outcome expectations, previous experience with SMBP grants, and supportive clinic leadership were identified as facilitators to implementation; Patients high social needs, SMBP-related Technology, and insufficient workforce and staff capacity were identified as barriers.

**Conclusion:** BT SMBP among FQHC patients is promising but challenges in integrating SMBP data into clinic workflow, workforce capacity to support the high social needs of participants and to assist in reacting to the more frequent BP data remain to be overcome.

## Introduction

Hypertension is the most important modifiable risk factor for cardiovascular disease, stroke, and Alzheimer’s Disease and Alzheimer’s Disease Related Dementias (AD/ADRD). [1-5] Currently, the US is facing a HTN epidemic; Nearly half of all Americans (47% or 116 million people) experience HTN, and only about half of Americans with HTN have controlled blood pressure (BP). [6-8] HTN disproportionately affects Black Americans; Black Americans have one of the highest prevalence of HTN in the world and are less likely than White Americans to have their BP controlled. [8] Disparities in BP are also evident among low-income Americans, who have higher prevalence, less awareness, less treatment, and less BP control than other Americans.[9]

New strategies to reach and support the control of BP in historically marginalized populations are needed. One approach to reach historically marginalized populations is through partnerships with Federally Qualified Health Centers (FQHCs). FQHCs provide comprehensive primary care to nearly 30 million Americans, 91% are low-income, and 60% are racial/ethnic minorities. [10-11] A complementary strategy to support BP control is via mobile health (mHealth), which leverages cellphone and smartphone devices already adopted by the majority of U.S. adults, and with high penetration among low-income and minoritized populations. [12]

In this context, the Health Resources and Services Administration (HRSA) and the Office of Minority Health partnered on the National HTN Control Initiative: Addressing Disparities among Racial and Ethnic Minority Populations (HTN Initiative). [13-14] Through this initiative, in 2021, HRSA awarded nearly $90 million in supplemental funding to FQHCs with a large proportion of patients with uncontrolled BP (<58.9%). The HTN Initiative’s goal is to enhance BP control using Bluetooth-enabled Self Measured Blood Pressure monitoring (BT SMBP) for patients with HTN and use their data to inform HTN treatment. [13] SMBP is the regular measurement of BP by the patient outside the clinical setting and is an evidence-based practice to lower BP and improve BP control. [15] SMBP is recommended to confirm the diagnosis of HTN, evaluate BP in response to treatment, and empower patients in managing their own BP, including antihypertensive medication adherence. [16-18] With the support of BT SMBP devices, combined with supporting mHealth cell phone apps, individuals can monitor their BP and communicate that data back to their healthcare providers in real or near real-time. We sought to understand the experience of Michigan-based FQHCs in implementing the HTN initiative via qualitative interviews based on the Tailored Implementation of Chronic Disease (TICD) Framework.

## Methods

This is a qualitative study using semi-structured interviews to assess barriers and facilitators to implementing Bluetooth-enabled Self Measured Blood Pressure monitoring (BT SMBP) for patients with HTN and the clinical use of their BP data to inform HTN treatment at FQHCs.

### Theoretical framework

The semi-structured interview guide was developed using the Tailored Implementation for Chronic Diseases (TICD) determinant framework to assess barriers and facilitators to BT SMBP patient initiation and clinic integration. The TICD is a comprehensive framework based on an extensive systematic review of determinants of practice in primary and secondary care as well as public health services. The TICD explores determinants of practice in seven domains which include guideline factors, health professional factors, patient factors, professional interactions, incentives and resources, capacity for organizational change, and social, political, and legal factors. [19] The interview guide began with a general description of the HTN initiative at each FQHC network. Open ended questions assessing all TICD domains except social, political, and legal factors were included in the interview guide. The interview guide concluded with a query of general advice for other FQHCs to implement BT-SMBP.

### Study Participants and procedures

Staff members from three Michigan-based FQHC Networks were invited to participate in semi-structured interviews from September to November 2022. As one of the FQHCs had not begun enrolling patients at the time of their interviews, a follow up interview was conducted with one staff member in February 2023. FQHCs were purposively selected based on 1) location in Michigan; 2) receipt of funding from the National HTN Control Initiative: Addressing Disparities among Racial and Ethnic Minority Populations; and 3) varied implementation strategies. Administrators at an FQHC with a longstanding partnership with the research team aided in connecting the research team with other FQHCs via email. Participants were compensated $20.

FQHC networks 1 and 2 have clinics located across the Detroit metropolitan area, including neighborhoods with significant Hispanic and Middle Eastern/North African populations. FQHC network 3 is located in central Michigan with clinics in a predominately Black community.

### Data Collection

Most interviews were conducted by AH (a woman MPH/researcher with experience implementing interventions in FQHCs and qualitative research experience) with support from CW (a woman with a BA in community health with extensive experience implementing interventions in under-resourced settings). Interviews were conducted by AH and CW, when possible, to allow for one researcher to take field notes. Interviews were conducted in-person at the FQHCs and were 15-60 minutes in length. Interviews were audio recorded, professionally transcribed, and reviewed by the research team for accuracy.

## Data Analysis

The research team created a codebook based on the TICD framework; Transcripts were deductively coded based on the TICD framework using Atlas.TI, a qualitative data analysis software. AH, CW, and MM (a woman with a BA with training in qualitative methods) independently coded two transcripts, meeting via Zoom after coding each transcript to discuss discrepancies until a consensus was reached. Edits to the codebook, including tailoring of definitions, examples, and notes, were made in real time in a web-based shared document. The remainder of the transcripts were randomly assigned to AH, CW, and MM to code independently.

AH exported coded data from Atlas.TI into an Excel spreadsheet for review. Data was sorted according to preliminary themes and refined into final themes. Final themes were reviewed and approved by CW and MM. Multiple research team members had full access to all the data in this study; The research team takes responsibility for the integrity of the data collection process, data analysis, and results.

## Results

### Participant Characteristics

Ten staff members from 3 Michigan-based FQHCs participated in interviews (FQHC 1: n=6, FQHC 2: n=1, FQHC 3: n=3). The number of interviews conducted at each FQHC was determined by ability to establish contact with FQHC staff, staff availability, and relevant staff given each FQHCs unique implementation status. Participants had a mean age of 43 years, half identified as White and were divided between clinic administration and direct patient care providers. Given that only one staff member from FQHC 2 participated in the interview, participant characteristics are presented in aggregate in Table 1.

**Table 1.** Participant Characteristics.

| Characteristic | Participants<br>n=10<br>n% |
| --- | --- |
| Age (Years) Mean (SD) | 43 (16.47) |
| Race |  |
| White/European American | 5 (50) |
| Black/African American | 2 (20) |
| Other | 3 (30) |
| Position |  |
| Administrative | 6 (60) |
| Care provider (CHW, health educator, RN)* | 4 (40) |

\* CHW= Community health worker; RN= Registered nurse.

### Overview of BT-SMBP Program by FQHC

FQHC 1 began implementation of BT-SMBP in February 2021 and created a large-scale, data-driven, CHW-supported remote monitoring program. Patients with uncontrolled BP in the prior year were identified in the electronic medical record (EMR) and contacted by the clinic’s CHW for enrollment. After 20 months, FQHC 1 enrolled over 580 patients into the program. As necessary, patients were offered the alternative to utilize paper BP logs. BP data was manually transferred from smartphone application (or paper BP log) to the clinic’s EMR by CHWs. Patients graduated from the program after two months of controlled BP. Participants were able to keep their BP device after graduation. FQHC 2 began implementation in September 2022 and structured the HTN initiative as a small-scale, short-term BP device loan program; Thirty-six BP devices were available to patients for one-month loans. Patients were identified in-real time during clinic appointments. FQHC 2 manually transferred patient’s BP data from the patient’s app to the clinic’s EMR. FQHC 2 had just begun enrollment, thus complete enrollment data was not available at the time of the interview. After an unsuccessful attempt to launch the BT-SMBP program internally, FQHC 3 worked with an outside company to launch a large-scale, social needs driven BT-SMBP program. The company was responsible for patient outreach, enrollment, and tracking BP measurements. Eligible patients were primarily identified via EMR searches; Patients with comorbidities and high social and economic needs were prioritized for enrollment. Participants were loaned a BT-BP device for the duration of their participation; The partner company facilitated data transfer directly into the clinic EMR, which was then accessible by providers. After their first two months, approximately 250 patients had enrolled. FQHC 3 intended to engage participants for at least a year.

### Barriers and Facilitators to implementing BT-SMBP in FQHCs

The identified barriers and facilitators to implementing the BT-SMBP program fell within 5 of the 7 TICD domains; These domains include individual health professional factors, capacity for organizational change, patient factors, guideline factors, and incentives and resources. We summarize our findings according to these TICD domains with illustrious quotes as evidence of each.

### Individual Health Professional Factors

#### Attitude/Outcome Expectancies

Facilitators were abundant in the Individual Health Professional Factors domain. Staff at all three FQHCs expressed overall positive attitudes towards the SMBP Program. Staff attitudes at FQHC 1 and 3 were tied to observed or anticipated patient outcomes including controlled BP, reduced emergency department visits, greater medication adherence, increased knowledge about hypertension, appointment attendance, and increased trust in the healthcare system, *“I think it’s going very well […] we took a whole population of patients that was listed as uncontrolled in 2020, and at the end of 2021, 58% of those [patients] was controlled.” (FQHC 1)* Staff at FQHC 1 used a variety of positive adjectives to express their appraisal of the program, including amazing, rewarding, successful, and worth it, *“[…] it’s very rewarding to see that their blood pressure is under control, it makes me happy just to feel that I had made a difference, you know, and we try as much as we can. Sometimes, like I said, there’s barriers, there’s hiccups, but it’s worth it. […] It’s worth it” (FQHC 1)* Staff at FQHC 3’s positive attitudes were also tied to anticipated clinician benefit, including the ability to create more tailored hypertension treatment plans, increased use of self-management goals in addition to BP medication, and alleviate their mental burden of worrying about a patient that presented with high BP, *“[The initiative will] help them not have to remember that patient that had that high blood pressure reading and did they return […] They know there’s that company out there that’s going to be collecting and letting us know when they don’t take a reading.”* FQHC 2 implemented a simplified version of SMBP. As they put it, *“Remote monitoring is when there is, like, a continuous flow of-of their blood pressure readings and data into the system. […] We do not have remote monitoring. So, we do the SMBP […] so basically, they just have to bring their phone with them to the return visit. They can […] show you the phone and-and the blood pressure readings on the phone, or on the app, rather*.” FQHC 2’s positive attitude towards the program was tied to their experience implementing this simplified program structure, “*I really thought it was going to be more complicated, but it wasn’t. Maybe if we were doing remote monitoring, that’s when it would get, like, a little bit more complicated.” (FQHC 2)*

#### Knowledge/Skills

Knowledge and skills also served as significant facilitators. FQHCs 1 and 2 shared that being a part of this and other hypertension grant initiatives helped prepare them to participate in the HTN initiative, for example, by helping them establish a workflow, (*“Like, we are actually part of five other hypertension programs. So, we’ve gotten the whole workflow down. Like, we have a team. We know what the barriers were, and how we addressed them.” FQHC 1)* and provided a protocol and foundation of knowledge, *(“I do work with Million Hearts, so […] that was very helpful in terms of, um, just giving a step-by-step guide to, you know, how to create and build this program and what steps do we need to check off, kind of, in order to have a well-functioning program. Also, information on different devices that are validated, which made it much, much simpler to choose a device that works for us.” FQHC 2)*.

#### Capacity for Organizational Change–leadership support

The capacity for organizational change domain had many facilitators. FQHC 1 and 2 expressed strong program leadership. At FQHC 1, staff shared providing CHWs with significant training opportunities and mentorship, including pairing new CHWs with seasoned CHWs and support from a health educator, *“I was actually hired in for the [HTN Initiative] and I actually learned my way up with the patients themselves because of our health educator, [name-redacted], who taught me to teach the patients. […]”* (FQHC 1). FQHC 2 shared hiring a staff member to oversee the program, *“So, I was hired on to fill a role within the [HTN Initiative]. […] So, I do most of the work regarding SMBP in terms of creating the program, implementation, training, finding, you know, which device would work best for our needs.”* Staff at FQHC 1 also mentioned that the support of their chief medical officer aided in provider program uptake, *“[…] our CMO is definitely a champion of it. Like he got us a lot of provider buy-in, and that really helped, like, new providers have to be on board to be able to, you know, identify the patients who are having symptoms, make that connection with the CHWs.”*

### Patient Factors

#### Patient motivation

At FQHC 1, staff widely cited that giving patients a free BP cuff motivated patient enrollment, and that social support from CHWs motivated program engagement. One staff member found that social support is especially appreciated by patients in light of the isolation caused by the COVID-19 pandemic, *“The patients, like, are so excited when [CHWs] call […] It’s like you see that they are appreciating having someone there as a support system to help them manage their BP. […] COVID was very isolating for many people. […] So, having somebody who is actually checking up on you and calling you, to kind of you know, ensure that you’re, like, staying healthy is really important for our patients.” (FQHC 1)*.

#### Patient beliefs/knowledge

A couple of staff at FQHC 1 and 3 identified patient education on hypertension as a facilitator to patient engagement, emphasizing the link between knowledge and self-efficacy, *“I think certainly from the patient standpoint, it is really a question of making sure that the patient is comfortable with recording his or her blood pressure, you know, at home, away from the clinic like that. Getting them used to the fact that there is a right way to measure the blood pressure and take the blood pressure. And I think with appropriate instruction, uh you know, on that particular…with our community health workers, our providers in the clinic, our support staff, to encourage them that can happen.”* (FQHC 3).

#### Patient needs

FQHC 1 cited patients’ lack of digital resources and digital literacy as barriers to enrollment, *“I think one of the reasons [patients decline to participate] is, it is, like, our digital literacy in our patients too, right? […] So, it’s sometimes hard for them […] they don’t have a smartphone. Like, […] it’s hard to kind of tell…the whole set up, once they don’t have all the right tools to be able to, you know, set it up. So, a lot of patients who don’t have one, we give them a physical paper log to document, but at the same time, it’s…well, there’s no point in having [BT SMBP] if you’re not going to be using your phone to integrate it and send that information back to your provider.”* More generally, FQHC 1 cited their patient population’s high level of social needs as a barrier to program engagement, *“ […] you’re going to deal with patients who have issues. You’re going to deal with patients who are mentally unsound, physically unsound […] you need to figure out what’s going on with them, and you have to dig deep with a patient. What are your barriers? Financial barriers […] literacy barrier[s]. They don’t know where they need to go. It’s housing barriers that we deal with, transportation barriers, and are you going to take the time to find the resources for your patient so they can get to that goal. Because yes, this is a blood pressure [program], but all the goals…other aspects associated with recovery of the health leads to that.”* In response to their patients social needs, several staff expressed that providing hypertension-related wrap-around resources, including sodium-free herbs, low-sodium cookbooks and hypertension resource guides, as potential facilitators to patient engagement, *“I’d like to have like maybe a little gift basket for them with, um, salt-free seasonings, certain herbs, things like that that they could use to cook because, um, it can be expensive and I had a lot of patients say, you know, Mrs. Dash is not cheap, um, those herbs are not cheap. […] I would like to see maybe some funding going towards like helping us get like little…do little packets for them and give it to them once we get…here’s your blood pressure, um, here’s a starter kit for you […].”*

### Guideline Factors

#### Feasibility

Technology-related barriers challenge program feasibility, from setting the patient up to getting the BP data to clinicians. FQHC 2 shared that the WIFI connection at one of their clinics was not strong enough to set up the patients’ devices, which has implications for conducting in-person patient training, *“[…] better internet connection, for one, would be a great resource, right. […] it makes all the difference if we can have the patient, that day, make sure that the device is paired, that that part is set up, that then they can show us how to do it, and we know they know how to do it. […] It also takes away some of the burden of having to, you know, then be with them on the phone, ‘I don’t know how to pair this device. […] I’m having issues,’ and you have to walk them through it. So, that lessens the burden there, if you can just get it done when they’re in clinic, right?”* Staff members at FQHC 1 shared problems pairing the

BP devices with cell phones. One staff member shared that many patients had a cellphone model that would not pair with the BP device’s corresponding smartphone application, *“Some of the phones don’t pair with the app. […] A lot of our patients come in with a certain type of Android that may not pair or something of the sort […]”* Another staff member shared that larger cuff attachments produced error messages, *“Another problem is in the blood pressure cuff device itself for this specific brand, the Welch Allyn one, because changing your cuffs, it can be smaller or larger and for the larger ones, whenever a patient needs to tighten it on, it doesn’t read it as fully tightened on properly, so it’ll constantly show an error message. So, sometimes I feel like I’m tightening the cuff too much on the patient just so that way they could get a reading when the average cuff puts it on, gives you a reading even if it’s too tight or too loose.”* Staff cited difficulty maintaining the pairing between participants BP devices and their cellphones, as well as staying logged into the corresponding app, “*A common problem with me [is] when a patient upgrades their phone […] and they forget to download the app. They’ll start taking their blood pressure and they’ll think that the device is automatically sending it to their phone. […] But when a patient upgrades phones, they think it’s still the same thing, but they don’t have the app it’s not connected via Bluetooth, so they’re just taking their blood pressure and thinking everything’s perfectly fine [technology-wise]”*. This disconnect places additional burden on clinic staff to contact the participant and re-pair the device, *“Sometimes, um, if they’re not logged into their phone, we can’t see the readings. So, that’s when we call and say, hey, are you logging into the app? They’ll so no. So, when they log back into the app and they take their blood pressure, all the other readings tend to just trend in.”*

Without the aid of a tailored software, staff at FQHC 1 and 2 shared finding makeshift ways to use existing computer software to track program participants devices and participation, including within their EMR, *“So, in Athena [EHR system], we place an order for loaning out a blood pressure monitor. […] We leave the order open in Athena […] you can leave the order open as an outstanding order which makes it helpful for tracking the devices, because then you don’t close it until the device is returned […] So, it-it just comes with that tracking aspect.”* (FQHC 2) These work-arounds do not alert staff if patients stop sending their BP, thereby allowing for periods of lapsed monitoring, *“I have [an] Excel sheet that I use, and I follow up with them on a monthly basis. […] I go in and look to see if they are still monitoring their blood pressure. If they’re not, ‘Hey! I just want to remind you to monitor your blood pressure. I’m looking at your readings right now, and I noticed that they haven’t been done in a couple of days. If you can please assist me and monitor them at least two to three times per week, that would give us the information that we need to ensure that we’re treating your diagnosis properly.’”* (FQHC 1). FQHC 3 also expressed similar technology challenges prior to switching to the assistance of a contracted partner, *“And having the technology to track each blood pressure cuff that went out and be able to track the patients we gave it to and then track who did and did not want to be enrolled in the system…in the program. And it was just too burdensome for the staff that we had, you know, at our sites. […] It was too burdensome. Yeah.”*

At FQHC 1 and 2, staff shared having to manually transfer participants BP data to the EMR, regardless of if the participant was connected to the BP device app or recording their BPs on a paper log, “*[BP readings don’t] automatically transfer over. So, I would need to go into one app to then print out all their blood pressure readings to then use our Athena [EMR] scan to upload it [pictures] to their chart.” (FQHC 1),* and *“[BPs] will be documented in the EMR once we get those […] if it’s a long list, right, they don’t have to input every single one, but they can put averages, right, or ranges are fine.” (FQHC 2).* In contrast, through FQHC 3’s remote monitoring partner, BPs flow directly from the patient into the patients EMR; This data is available to clinicians in multiple formats, *“It is up to the provider. They can always go to the system and look at each measurement…but coming into the EMR automatically is a monthly average reading that’s going to a flow sheet so they can look at that graphic, if they choose.”*

#### Incentives and Resources

Incentives and resources were dominated by barriers. Human resources, including staffing and staff capacity, were barriers to implementation. FQHC 1 shared having high CHW turnover rates, disrupting the continuity of the program and the social relationships made between CHWs and their patients, “*Well, I think we had high staff turnover […] So, that kind of made it difficult, because you would train a CHW or staff would take over the hypertension grant and then they would leave after four to six months […] and then we would have to do the process again which it was just like, you know, like the patients want to see familiar faces too.” (FQHC 1).* Insufficient pay motivated staff turnover, *“I think this group [CHWs] that we have now is sound. I’m happy with them, and I think they do it because they like the work and not for the check.”* FQHC 3 stated being heavily impacted by the industry-wide workforce shortage; They shared that the shortage is experienced differently in the FQHC setting, given the role they fill and the suite of services they provide, *“[…] from a staff perspective, I think our largest issue is, like I said, at the beginning, is just workforce shortages. […] People have historically always been spread thin, and FQHCs wear a lot of hats. But I mean, COVID has had its impact on the healthcare workforce. That’s really our biggest constraint with anything.”*

Staff at the FQHCs shared already having limited capacity. At FQHC 1, CHWs shared juggling a myriad of health education and screening responsibilities which make balancing their workload with the SMBP program difficult, “*As of right now, I’m starting to focus on A1C checks, so, diabetic management. I also help out with [social determinants of health]. I’m doing tobacco screening. I’m doing BMIs, health education, prediabetes. So, being a CHW is almost like being a jack of all trades. […] So, trying to focus on one patient for one certain thing and then having to jump back to hypertension, it gets a little bit difficult once you get a larger workflow […]”*. The dysergy of the workforce shortage and limited workforce capacity resulted in FQHC 3 changing from an mainly clinic-run program to contracting a remote monitoring company, “*So, I mean, everyone in healthcare is frankly understaffed and overburdened […] So then when you add on another initiative like this where we’re looking at patients, we’re engaging with them, educating them, enrolling them, monitoring them, and so on, that’s…that can be a big lift. So, what we’ve done is we found a partner that will not only help us to reach out to and enroll the patients but will help us with the 24/7 monitoring. […] it’s a great fit for our capacity, […] what we do is we pull the lists.”*

FQHCs shared incongruence between program funding and objectives, as one staff member at FQHC 3 put it, *“when I first looked at it, my analysis of the funding that was made available through the [HTN Initiative] and the number of patients they wanted engaged, I mean there was a strong disconnect […] in terms of what they were giving and what the expectation was. […] I would like to know, I mean, if there was [an FQHC] who just really knocked it out of the park, not only how did they approach it, but how did they fund it? Because my hunch is they definitely had to braid other funds in there to make it happen.”* Limited grant funding resulted in program structures with diminished patient reach and duration of engagement; Both FQHC 2 and 3 acknowledged their inability to bill insurance exacerbated their funding challenges. FQHC 2 shared that the funding was neither sufficient to finance the assistance of a remote monitoring company nor the purchase of BP devices for the number of patients the grant wanted them to reach, *“We were looking at [Remote monitoring company] for a while. […] I don’t think that they accepted Medicaid and that was an issue in terms of whether we could afford, because that would…the insurance would at least pay for some of, you know, their services. So then, more of the burden then would fall on the clinic. […] We do have a lot of patients on Medicaid or no insurance, right.”* As a result, FQHC 2 created a BP device loan program with a limited number of devices (12) for brief periods of engagement (1 month), “*Um. So, we figured a loaning program would be best, so that’s what we use.[…]The device can be loaned out for up to one month […] We figured one month, because that gives enough time, if the doctor really wants a nice length of time for home measurements, but not so far out that the patient is more likely to just forget they have the monitor and not return it. [Laughing] Right?”*. FQHC 3 shared that their remote monitoring company priced according to the number of patients being monitored, which limited their reach, *“Because of the costs, you know, our hands are tied at how many monitors can go out and how many pieces of equipment can be monitored at a single time. […] If we were able […] to bill out to the insurance company, the cost of the equipment plus the monthly monitoring fee, um, and where the provider is just giving a care plan and treatment, you know, indications or medication adherence […] you could certainly get more on your list.”*

**Table 2.** Barriers and Facilitators by Tailored Implementation in Chronic Disease (TICD) Domain.

| Domain | Barrier/Facilitator | Construct |
| --- | --- | --- |
| Individual Health<br>Professional Factors | + | Attitude |
|  | + | Expected outcomes |
|  | + | Domain Knowledge |
| Patient Factors | + | Patient motivation |
|  | + | Patient<br>beliefs/knowledge |
|  | - | Patient needs |
| Capacity for<br>Organizational<br>Change | + | Capable leadership |
|  | + | Strengths of<br>Supporters |
| Guideline Factors | - | Feasibility |
| Incentives and<br>Resources | - | Human Resources |
|  | - | Financial Resources |

**\*+ signifies facilitator; - signifies barrier.**

## Discussion

In summary, three Michigan-based FQHCs participated in semi-structured interviews about their experience implementing the HTN Initiative; Interviews were grounded in the TICD framework. Each FQHC was at a different stage of implementation, ranging from a year and a half of implementation to preparing to begin. Implementation strategies varied significantly by FQHC, ranging from a small, simplified, clinic-run loaner BT-SMBP program to a large-scale, outsourced model with continuous BP data transfer.

Facilitators to program implementation were abundant in the individual health professional factors and capacity for organizational change domains. Across FQHCs, staff demonstrated positive attitudes towards the program; Many staff’s positive attitudes were tied to observed or anticipated patient health outcomes, and some staff’s attitudes were also tied to the anticipated benefits for clinicians. This is notable particularly given their multiple other responsibilities and the increased workload of BT-SMBP, FQHC staff remained positive about the program, highlighting their commitment to their patients’ health and wellbeing. Knowledge and skills acquired through participating in other SMBP initiatives provided the FQHCs with a foundation to build their programs. High capacity for organizational change, including dedicated program leadership and mentors, training opportunities, and provider champions aided implementation. Some facilitators were also present in the patient factors domain. BP devices that participants could keep and social support from CHWs were identified as strong patient motivators to program adoption and engagement.

Implementation faced several barriers; Barriers were prevalent across the patient factors, guideline factors, and incentives and resources domains. Unmet social needs among patients were identified as competing priorities. Patients’ lack of digital resources and low digital literacy deterred patient enrollment and engagement. Technology posed a significant barrier to program feasibility as a common cellphone model did not pair with the BP device, devices produced error messages, and patients struggled to maintain the pairing between their phone and the BP device. FQHCs also shared not having the technological infrastructure to track loaned BP devices, track patient engagement, and manage SMBP data. Such disjointed and piecemeal technology resulted in increased staff burden, requiring manual BP data transfer by CHWs into patient’s EMRs. Short staffing and overburdened staff, ability to implement large clinic run programs, as staff were unable to identify, enroll, and engage patients in the program in addition to their existing workload. FQHCs managed this by scaling back their program or outsourcing initiative enrollment and data transfer. Only once clinics have the necessary technology and designated program staff can they truly engage in real time monitoring. Overall, insufficient funding negatively impacted clinics reach and duration of engagement, made contracted help financially inaccessible, and resulted in BP device loan-based programs. Such short durations of participant engagement may result in insufficient BP data to inform provider clinical decisions. From a patient standpoint, the engagement period may be too brief to cultivate knowledge of their BP and the efficacy of BP medications, thereby decreasing the benefit of increased medication adherence.

The identified barriers and facilitators present several pathways of action to bolster program implementation. FQHCs with experience implementing SMBP-related initiatives can serve as a resource to less experienced FQHCs. This information could be conveyed via learning health collaboratives, clinic champions or shared protocols. Financially accessible technology that can aid in tracking patient engagement and convey patient data directly into the EMR in clinically useful way has the potential to overcome several identified barriers and continue to support staff attitudes towards the program.

This study has limitations. This study represents a small sample of 3 FQHCs in a limited geographic area across southeast and central Michigan. The results from these interviews may not capture the barriers and facilitators to a breadth of FQHCs. Moreover, across these 3 FQHCs, we had a small sample size of 10 participants, which did not include physicians due to scheduling challenges nor patients. Physicians may have additional valuable insights as to the barriers and facilitators to accessing and utilizing patient BP data to inform clinical decisions. Similarly, this initiative was implemented across clinics that serve racially and ethnically diverse patients. Patients, including participants and those who declined, may provide, for example, insights into needed cultural and linguistic tailoring to support patient enrollment and engagement. Lastly, as this was the research team’s first interaction with two of the three FQHCs, staff may have provided socially desirable answers in effort to positively represent their clinics and their experience implementing this program. However, this risk was attenuated by conducting 1-on-1 interviews by researchers knowledgeable about the FQHC environment and qualitative methods.

FQHCs took varied approaches to implementing the HRSA SMBP program. Across clinics, the individual health professional factors, capacity for organizational change, and patient factors domains contained facilitators to implementation. The guideline factors and incentives and resources domains were dominated by barriers. Overall, BT-SMBP among FQHC patients is promising but challenges remain to be overcome.

## Data Availability

Data is available upon request.

## Abbreviations

HTN: Hypertension
FQHC: Federally Qualified Health Center
HRSA: Health Resources and Services Administration
BP: Blood pressure
BT SMBP: Bluetooth-enabled self measured blood pressure
SMBP: self measured blood pressure
AD/ADRD: Alzheimer’s Disease and Alzheimer’s Disease Related Dementias
TICD Framework: Tailored Implementation of Chronic Disease Framework
EMR: Electronic medical record
CHW: Community health worker

## Acknowledgements

All co-authors have reviewed this publication and accept their authorship.

## Supplemental Material

